# Atypical B cells and impaired SARS-CoV-2 neutralisation following booster vaccination in the elderly

**DOI:** 10.1101/2022.10.13.22281024

**Authors:** Isabella A.T.M. Ferreira, Colin Y.C. Lee, William Foster, Adam Abdullahi, Zewen Kelvin Tuong, Benjamin J Stewart, John R. Ferdinand, Stephane Guillaume, Martin O.P. Potts, Marianne Perera, Benjamin A. Krishna, Ana P. Alonso, Mia Cabantous, Steven A. Kemp, Lourdes Ceron-Gutierrez, Soraya Ebrahimi, The CITIID-NIHR BioResource COVID-19 Collaboration, Paul Lyons, Kenneth GC Smith, John Bradley, Dami A. Collier, Sarah A. Teichmann, Laura E. McCoy, Paul A. MacAry, Rainer Doffinger, Mark R. Wills, Michelle Linterman, Menna R. Clatworthy, Ravindra K. Gupta

## Abstract

Age is a major risk factor for hospitalization and death after SARS-CoV-2 infection, even in vaccinees. Suboptimal responses to a primary vaccination course have been reported in the elderly, but there is little information regarding the impact of age on responses to booster third doses. Here we show that individuals 70 or older who received a primary two dose schedule with AZD1222 and booster third dose with mRNA vaccine achieved significantly lower neutralizing antibody responses against SARS-CoV-2 spike pseudotyped virus compared to those younger than 70. One month after the booster neither the concentration of serum binding anti spike IgG antibody, nor the frequency of spike-specific B cells showed differences by age grouping. However, the impaired neutralization potency and breadth post-third dose in the elderly was associated with enrichment of circulating “atypical” spike-specific B cells expressing CD11c and FCRL5. Single cell RNA sequencing confirmed an expansion of *TBX21-, ITGAX*-expressing B cells in the elderly that enriched for B cell activation/receptor signalling pathway genes. Importantly we also observed impaired T cell responses to SARS-CoV-2 spike peptides in the elderly post-booster, both in terms of IFNgamma and IL2 secretion, as well as a decrease in T cell receptor signalling pathway genes. This expansion of atypical B cells and impaired T cell responses may contribute to the generation of less affinity-matured antibodies, with lower neutralizing capacity post-third dose in the elderly. Altogether, our data reveal the extent and potential mechanistic underpinning of the impaired vaccine responses present in the elderly after a booster dose, contributing to their increased susceptibility to COVID-19 infection.

## Introduction

The adenovirus vectored AZD1222 vaccine (ChAdOx1 nCov-19) was one of the first vaccines approved for use in the United Kingdom in early 2021^1^, and came shortly after rollout of the mRNA BNT162b2 ^2^. During initial scale up of vaccination in early 2021, there were several variants of concern circulating, including Alpha (B.1.1.7) and Beta (B.1.351); vaccines were shown to confer protection to Alpha but not Beta ^3-5^, likely due to escape from neutralizing antibodies mediated by the spike mutation E484K.

With emergence of the Delta variant ^6-8^ coupled with waning neutralizing antibodies ^9,10^ booster doses were recommended. Emergence of the Omicron BA.1 variant ^11^ further strengthened the argument for booster doses when data emerged showing broader neutralization compared to two doses ^12-14^. In contrast to neutralizing antibody titers, Spike-specific B cell frequencies remain stable across time and after third dose neutralizing antibodies appear more able to tolerate receptor binding domain (RBD) mutations, consistent with ongoing antibody maturation ^15-17^.

Long-lived B cell immunity, important in maintaining immunity elicited by vaccines ^17,18^, is affected by immune aging in the elderly and moreover, functional recall to SARS-CoV-2, is lower than in younger individuals ^19,20^. Our previous work indicated that age broadly affected immune responses in those vaccinated with the mRNA vaccine BNT162b2 ^21^, particularly following first SARS-CoV-2 vaccine dose. This difference diminished after the second dose of the vaccine, but the T cell response remained poorer in the elderly despite second mRNA vaccine dose.

Here, we aimed to determine the impact of age on responses to the third vaccine dose, and to understand the mechanistic underpinning of the differential immune responses observed with increasing age. In the UK, individuals vaccinated with AZD1222 either received the BNT162b2 or mRNA-1273 vaccine boosting approximately six months after their second dose ^22^. We focused on individuals who received two doses of AZD1222 and an mRNA booster vaccine because we and others have reported lower neutralizing antibody responses following two doses of AZD1222 compared to BNT162b2 ^6,12,23^. We measured the breadth and durability of vaccine-elicited neutralizing antibody and T cell responses across 36 individuals receiving AZD1222 as their primary two-dose course. We also applied multiparameter flow cytometry and single cell RNA sequencing to peripheral blood mononuclear cells (PBMCs) obtained one month following the second dose of AZD1222, and one month post-BNT162B2 booster dose, comparing cell phenotypes, single cell transcriptomes and antigen receptor sequences longitudinally across age groups.

## Results

### Binding and Neutralising antibody responses following two doses of AZD1222 and third dose mRNA vaccination

We enrolled 60 individuals who had been vaccinated with two doses of AZD1222 and one mRNA booster vaccine (either BNT162b2 or mRNA-1273). Blood draws were taken 1 month post second dose, 6 months post second dose, and 1 month post booster third dose (**Figure 1A, Supplementary Figure 1**). Thirty-six individuals had samples available for all time points and were N antibody negative at all time points. The median age of study participants was 67 years of age and no individuals were aged above 75 (**Supplementary Figure 1**). We initially measured SARS CoV-2 spike (S) total IgG along with N total IgG using Luminex based flow cytometric analysis ^24^, the latter to exclude any individuals who may have had SARS-CoV-2 infection from our study.

**Figure 1:**
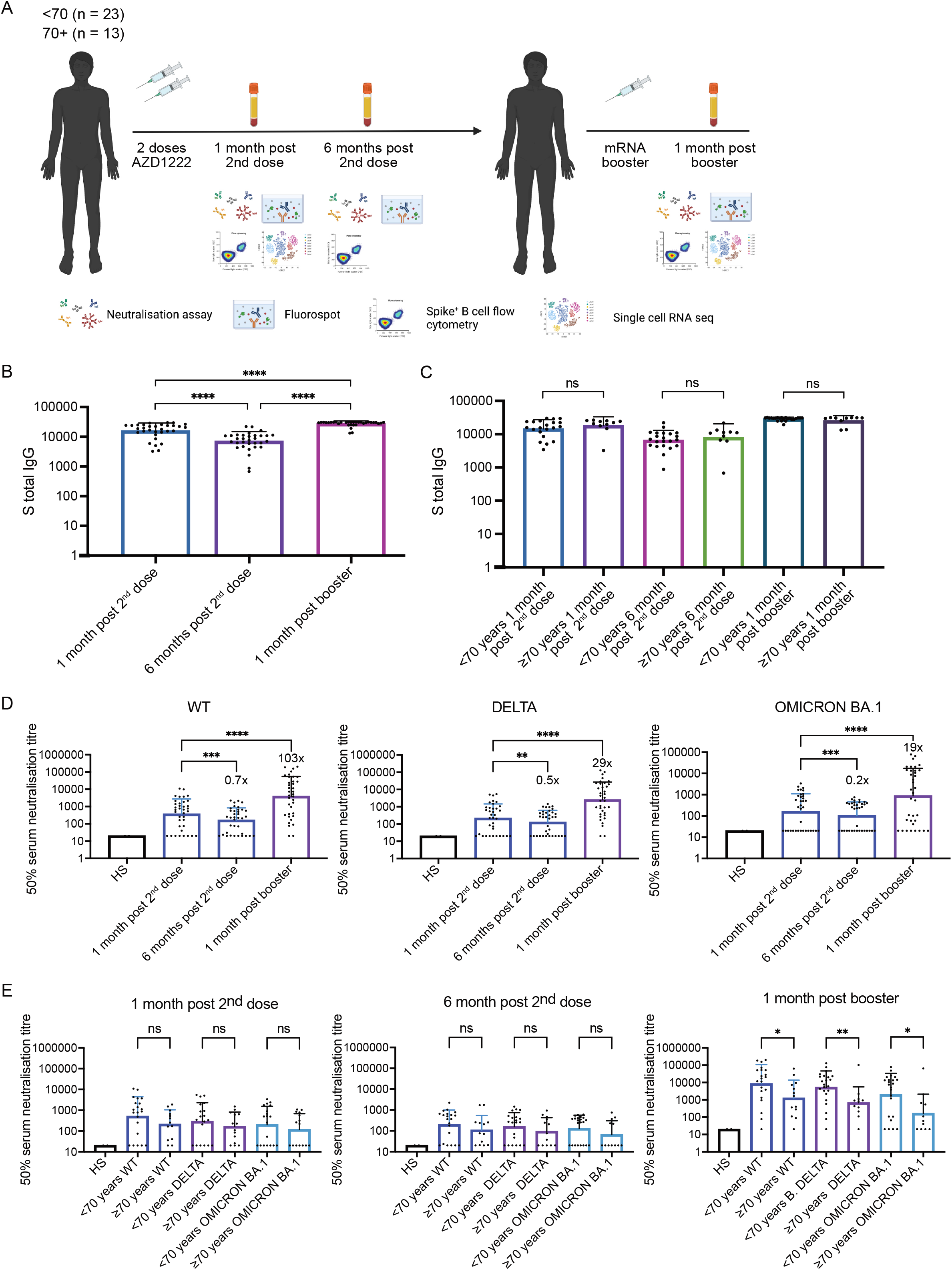
Longitudinal neutralising plasma antibody titres against Wu-1 D614G WT, Delta, and Omicron BA.1 variants from AZD1222 vaccinated individuals boosted with an mRNA-based vaccine. (A) Study design - 36 individuals vaccinated with AZD1222 and boosted with an mRNA-based vaccine were recruited. Longitudinal blood draws occurred 1 month post second dose, 6 months post second dose, and 1 month post booster. (B) Total anti-Spike IgG binding antibody responses at 1 month post second dose, 6 months post second dose, and 1 month post booster. Wilcoxon matched-pairs signed-rank test was used. **** p < 0.0001. (C) Total anti-Spike IgG binding antibody responses at 1 month post second dose, 6 months post second dose, and 1 month post booster stratified by those below age 70 and those age 70 and above. Mann-Whitney test was used. NS is non-significant. (D) Neutralisation titres (ID50) of sera was measured against Wu-1D614GWT, Delta, and Omicron for each time point. A Wilcoxon matched-pairs signed-rank test was used to determine significance in titres between time points. ** p < 0.01, *** p < 0.001, **** p < 0.0001. (E) Neutralisation titres (ID50) against against Wu-1 D614G WT, Delta, and Omicron BA.1 stratified by those below age 70 and those age 70 and above. Mann-Whitney test was used. NS is non-significant. * p < 0.05, ** p < 0.01.

Total spike (S) IgG, as measured by mean fluorescence intensity (MFI), decreased between 1 month and 6 months post-second dose AZD1222 (p < 0.0001), with a significant increase evident following the booster mRNA vaccination (p < 0.0001) (Figure 1B). A significant increase was also present when comparing 1 month post-second dose and 1 month post-booster (p < 0.0001) (**Figure 1B, Supplementary Figure 2**), indicating that the booster had an additive effect on S total IgG. When comparing <70 and ≥70 age groups, there was no significant difference in S total IgG at any timepoint (**Figure 1C**).

We assessed neutralizing antibodies using a previously developed spike-pseudotyped lentiviral neutralization assay^21^. SARS-CoV-2 D614G wild type (WT) spike was used as the comparator spike against the Delta and Omicron variants of concern. Overall, geometric mean titers (GMT) as a measure of the mean ID50 at each time point showed a decrease from one to 6 months post-second dose (GMT=368.9 (standard deviation (SD) of 6.44) and 143.6 (SD = 5.19) respectively), but a robust augmentation 1 month following the booster mRNA vaccine dose (GMT=3440 (SD = 15.40), **Figure 1D**).

Across the ancestral D614G, Delta and Omicron variants, there was a significant decrease in neutralizing antibodies 6 months post second dose compared to 1 month post second dose (P < 0.0001, p < 0.0011 and P < 0.0003 for D614G, Delta and Omicron respectively). Fold changes indicated relatively modest waning in circulating neutralizing antibodies against WT and Delta between 1 month post second dose and 6 months post second dose (**Figure 1D, Supplementary Figure 3**). A greater degree of waning was observed for Omicron (**Figure 1D**). Boosting with an mRNA-based vaccine showed a significant increase in neutralizing antibodies across the three variants compared to 1 month post second dose (103x fold increase between post-second dose and post-booster for WT, 29x fold increase Delta, and 19x fold increase for Omicron) (**Figure 1D**).

We next assessed the impact of age on boosting of neutralizing antibody responses. No differences in serum neutralizing antibody titers were observed across age groups for the time points of 1 month post second dose and 6 months post second dose across the three variants of concern (**Figure 1E**). As expected, there was a log decrease in neutralising antibody titers between 1 month post booster and 6 months post booster. However, the ≥70 group demonstrated significantly lower neutralizing antibody GMTs 1 month post booster (Delta: p < 0.0240; Omicron: p < 0.0450). After the mRNA booster vaccine 4% of individuals <70 years old were non-neutralisers (ID50 titers of <20) and 8% >70 year olds were non responders for WT. For Delta, 4% of <70 year olds were non-neutralisers and 15% of ≥70 compared to 17% of <70 for Omicron and 22% of ≥70 (**Figure 1E, Supplementary Figure 3**). In summary, the antibody measurements show that the mRNA booster elicits a robust augmentation in neutralizing antibodies, with a diminished response in participants aged 70 years or more.

### Single cell RNA sequencing identifies age-related differences in B cell vaccine response

We performed scRNAseq to assess gene expression, as well as single cell B cell receptor (BCR) and T cell receptor (TCR) sequencing, in PBMCs taken 1 month post dose 2 AZD1222 (n=20 participants) and 1 month post-mRNA booster (n=19 participants). Following the application of a rigorous quality control pipeline, 99,384 cells were available for analysis, and annotated using CellTypist ^25^ and canonical marker gene expression, identifying 15 major cell types, including CD4 and CD8 T cells, B cells, monocytes (classical and non-classical), classical dendritic cells (DCs), plasmacytoid DCs (pDC), natural killer (NK) cells, innate lymphoid cells (ILC), and mucosal-associated invariant T cells (MAIT) (**Figure 2A, Supplementary Figure 4)**.

**Figure 2:**
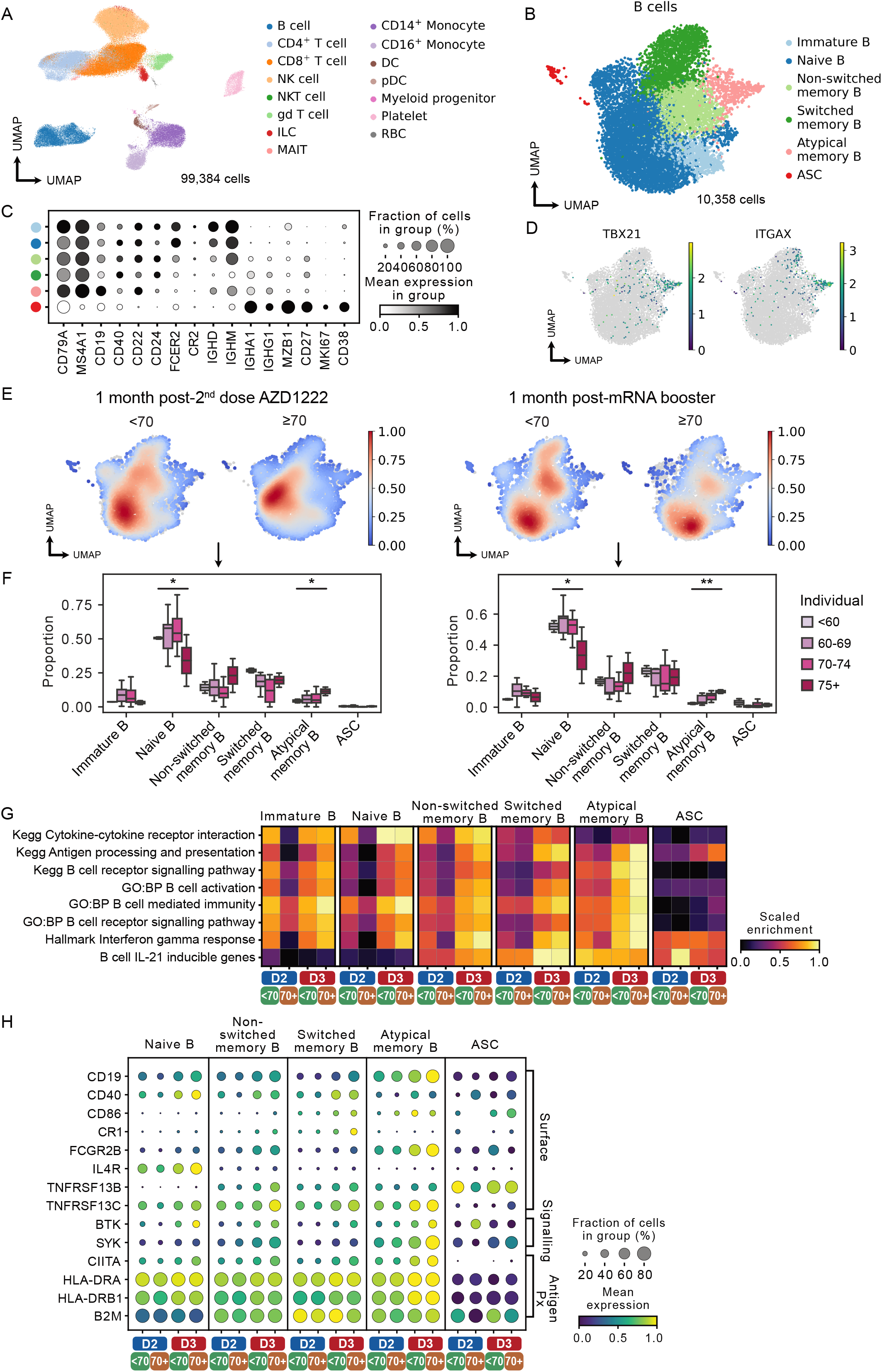
Single-cell RNAseq identifies age-associated differences in B cell responses post-vaccination. (A) Uniform manifold approximation projection (UMAP) of cell types identified by scRNAseq of PBMCs in samples taken 1 month post dose 2 AZD1222 (n=20 subjects) and 1 month post-mRNA booster (n=19 subjects). (B) UMAP of subsetted B cells annotated by canonical marker gene expression (C). (D) Atypical memory B cells express *TBX21* and *ITGAX*. (E) Density plots showing B cell abundance in <70 and 70+ individuals 1 month post dose 2 AZD1222 (left) and 1 month post-mRNA booster (right). (F) Proportions of B cell subsets 1 month post dose 2 AZD1222 (left) and 1 month post-mRNA booster (right), in individual study participants, in different age groups. Significance testing using Kruskal-Wallis one-way test. (G) Heatmap showing gene set expression in B cell subsets in <70 and 70+ individuals 1 month post dose 2 AZD1222 (D2) and 1 month post-mRNA booster (D3). (H) Selected differentially expressed genes driving differences in (G).

When considering the B cell compartment in isolation, fine clustering identified a small number of antibody-secreting cells (ASC), as well as immature, naïve, non-switched and switched memory B cells, and a population of *TBX1-* (encoding TBET) and *ITGAX*-(encoding CD11c) expressing ‘atypical’ memory B cells (also previously described as exhausted or age-associated B cells ^26^) (**Figure 2B-D, Supplementary Figure 5**). The abundance of naïve B cells was lowest in those ≥70 years of age, and there was an increase in atypical memory B cells with increasing age, both following dose 2 AZD1222 and mRNA booster vaccines (**Figure 2E-F**).

Analysis of geneset expression showed differences, both between vaccine doses and according to age; Overall, the magnitude of expression of several relevant genesets across B cells subsets was greater at 1 month following the mRNA booster (D3) compared with the same timepoint post-dose 2 AZD1222 (D2) (**Figure 2G**). This difference was particularly marked in antigen-experienced subsets (memory and atypical B cells), for example, ‘*antigen processing and presentation’* pathway genes such as *CD40*, were minimally expressed on these cell subsets post-D2 but demonstrated robust expression post-D3 (**Figure 2G-H, Supplementary Figure 5**). ‘*Cytokine-cytokine receptor interaction’* genes were also increased post-D3 compared with D2, particularly in naïve and non-switched memory B cells, and included *IL4R* and BAFF receptors *TNFRSF13B* (encoding TACI) and *TNFRSF13C* (encoding BAFF-R) (**Figure 2G-H**). ‘*Interferon gamma response*’ and ‘*IL-21 inducible genes*’ were also increased following D3, which are important in B cell expansion in the germinal centre and for class switch recombination^27^. Notably, the difference in geneset expression between D2 and D3 samples was more marked in the ≥70 age group; for example, in naïve B cells post-D2, ‘*Cytokine-cytokine receptor interaction’* genes showed modest expression in those aged <70, but were barely detectable in cells from participants aged ≥70. In contrast post-D3, this geneset was robustly expressed at similar levels in both age groups (**Figure 2G**). Indeed, in atypical B cells post-D3, expression of ‘*antigen processing and presentation’* and B cell activation pathway genes in the ≥70 age group actually exceeded that of younger subjects (**Figure 2G**). Altogether, our data suggests that individuals ≥70 years old have less B cell activation post-dose 2 AZD1222 than individuals <70 years old, but that this age-associated difference is no longer evident, or even reversed, after the mRNA booster dose, which leads to more robust and/or prolonged transcriptional changes in B cells in both age groups, despite the differences in neutralizing antibody titres observed between age groups (**Figure 1E**). To investigate this further, we next focused our analyses on B cells that are reactive to the SARS-CoV-2 Spike protein or its receptor binding domain (RBD)

### Virus-specific atypical B memory cells expanded in the elderly post-mRNA booster

We investigated the antigen-binding capacity of memory B cells in our scBCRseq data using their CDR3 sequences, since CDR3 plays a dominant role in antigen binding and specificity (Noel et al. 1996; Xu and Davis 2000). To do this, we used a public COVID-19 antibody database (Cov-AbDab) which has curated >10,000 CDR3 experimentally-validated sequences for SARS-CoV-2-specific antibodies ^28^. When considering paired heavy and light chain CDR3 sequences in our scBCRseq data, we identified n=152 matched BCRs in our memory B cell subsets with putative SARS-CoV-2 surface antigen-binding capacity (**Figure 3A**). SARS-CoV-2 binding BCRs were defined as cells with an identical light-chain CDR3 (CDR3-L) sequence and a paired heavy chain CDR3 (CDR3-H) with significant motif similarity to an antibody within the Cov-AbDab (**Supplementary Figure 6**). We also tested for motifs that were enriched in post-vaccination BCRs compared to control COVID-19-naïve B cells. Atypical B cells comprised a larger proportion of the memory cells expressing SARS-CoV-2 antibody-matched BCRs in subjects ≥70 years of age compared with those <70 years of age (**Supplementary Figure 6**), and this effect was particularly prominent following the mRNA booster vaccine (30% and 5% in ≥70 and <70 respectively, **Fig. 3B**).

**Figure 3:**
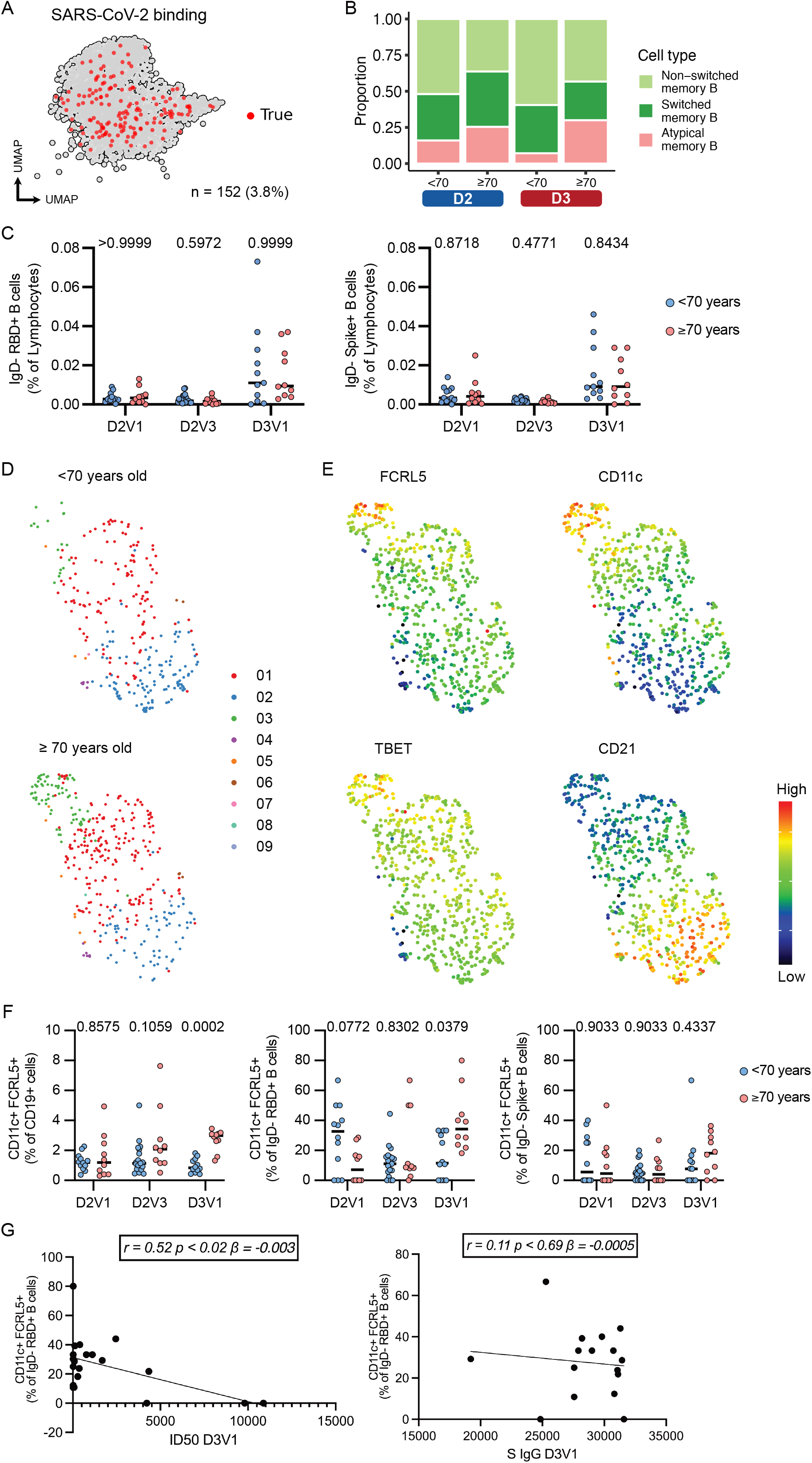
Older individuals have a higher frequency of antigen-specific atypical B cells after mRNA vaccine booster. (A) CDR3 heavy and light chain scBCR sequences with matched sequences or shared motifs with SARS-CoV-2 binding antibodies from Cov-AbDab. (B) Proportion of scBCR sequences in (A) expressed by atypical B cells, switched and non-switched memory B cells at each time point in <70 and 70+ age groups. (C) IgD-RBD+ and IgD-Spike+ B cell frequency, (as a % of live, single, lymphocytes) at each timepoint. (D) UMAP clustering analysis of a subset of IgD-RBD+ B cells from D3V1. (E) Relative MFI of indicated markers in UMAP clustering analysis from (D). (F) Atypical (CD11c+ FCRL5+) B cell frequency, (as a % of CD19+ cells, IgD-RBD+ and IgD-Spike+ cells respectively) at each timepoint. (D2V1 = 1-month post second dose, (D2V3 = 6 months post second dose, and D3V1 = 1-month post booster. For (C) and (F), each symbol represents a unique biological sample; multiple Mann-Whitney tests per row with Holm-Šidák multiple testing correction was used.

To validate these findings, we phenotypically assessed circulating SARS-CoV-2 RBD- and Spike-binding B cells by high-content spectral cytometry. Overall, there was an increase in the proportional representation of both RBD- and Spike-binding non-naïve (IgD-) B cells among lymphocytes 1 month post-mRNA vaccine booster, compared to 1 and 6 months post-second dose AZD1222, that was comparable in subjects <70 and ≥70 years of age (**Fig. 3C, Supplementary Figure 7**). However, unbiased machine learning analysis showed an altered distribution of IgD-Spike-binding B cell subsets between the <70 and ≥70 years old groups (Figure 3D). One such subpopulation expanded in the ≥70 years old group had increased expression of FcRL5, CD11c and TBET, with low expression of CD21 – consistent with an atypical memory B cell phenotype (**Figure 3E**). A distinct population of CD11c+ FCRL5+ atypical B cells was evident using conventional bi-axial gating (Supplementary Figure 7D-F), and expanded among subjects in the ≥70 age group, following the mRNA booster vaccine (mean of 0.975% vs 2.7% respectively p=0.0002 **Fig. 3D, F**). When considering RBD- and Spike-binding non-naïve B cells at this timepoint, there was also a greater proportion of antigen-specific atypical non-naïve B cells in older subjects compared to younger subjects, with an average of 39% of IgD-RBD+ B cells having atypical phenotype within the ≥70 group (p < 0.038 for RBD, **Figure 3F**).

In order to demonstrate that the presence of atypical cells was linked with poorer neutralization, we performed correlation analysis between atypical FcRL5, CD11c+ B cells as a proportion of IgD-RBD+ B cells and (i) binding anti-spike IgG titres and (ii) neutralization titres. We observed no relationship between atypical B cell abundance and serum IgG, but a significant negative correlation with serum neutralizing activity (**Figure 3G**). Taken together, our data suggests that the mRNA vaccine booster is able to support the expansion of vaccine-specific memory B cells, but that older age is associated with a skewed B cell differentiation towards atypical memory B cells that have been reported to be less effective at contributing to protective humoral immunity ^26,29,30^.

### T cell responses following two doses of AZD1222 and third dose mRNA vaccination

T cells are thought to maintain protection against SARS-CoV-2 infection when neutralizing antibody levels wane over time ^31^. We therefore considered the single cell T cell and innate lymphocyte transcriptomes in isolation, comprising 72,507 cells, including naïve, effector memory (EM), terminal effector (TE) and cytotoxic CD4 T cells and naïve, EM and TE CD8 T cells, as well as CD16+ and CD56+CD16-NK cells, ILCs, MAITs, NKT and □ □T cells (**Figure 4A-B**). There was a marked increase in abundance of TE CD8 T cells with increasing age, both following dose 2 AZD1222 and mRNA booster vaccine (**Fig. 4C-D**).

**Figure 4:**
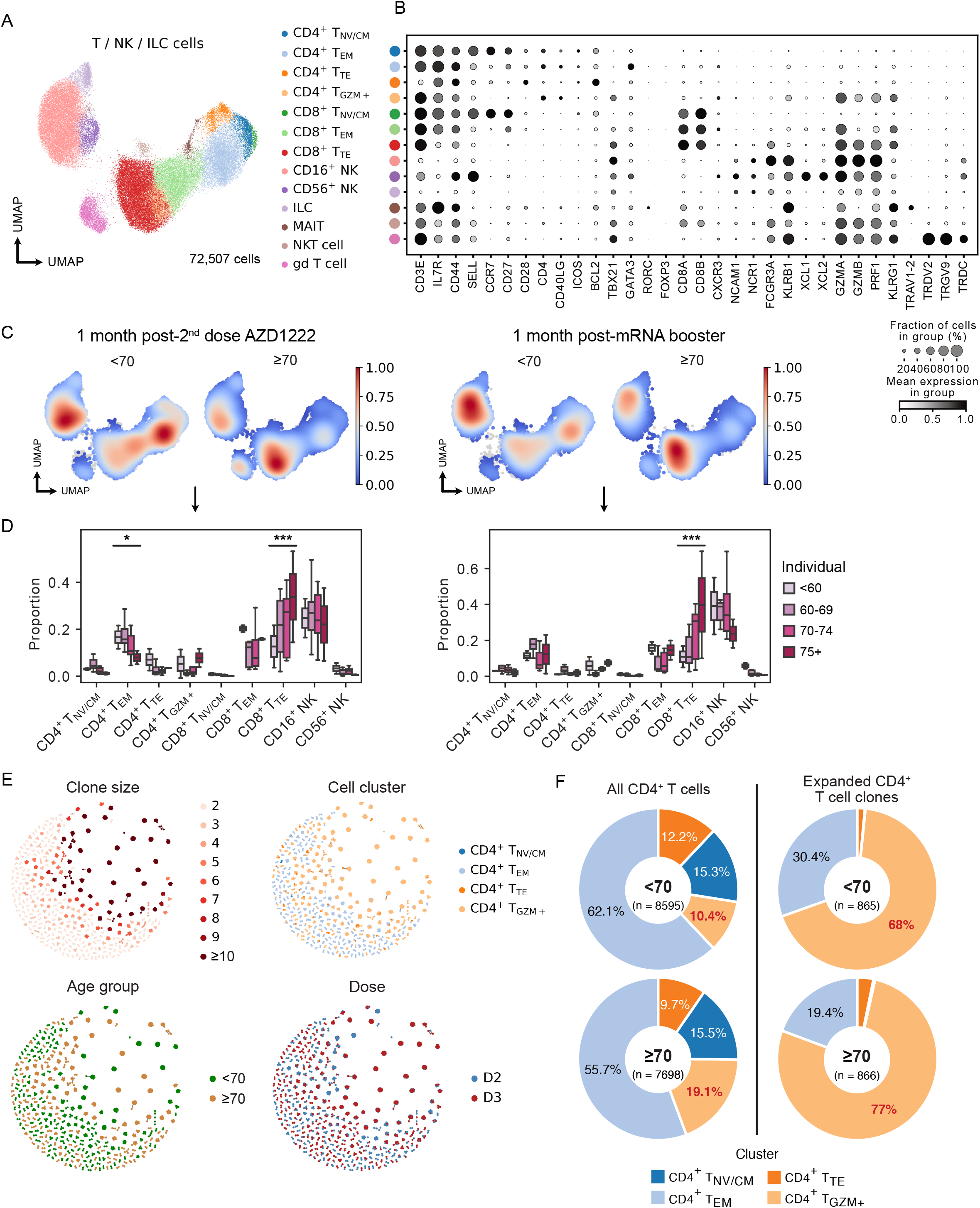
T cell responses to two doses of AZD1222 and an mRNA booster. (A) UMAP of subsetted T cells, natural killer (NK) an innate lymphoid cells (ILCs) annotated by canonical marker gene expression (B). (C) Density plots showing T/NK/ILC cell abundance in <70 and 70+ individuals 1 month post dose 2 AZD1222 (left) and 1 month post-mRNA booster (right). (D) Proportions of T/NK/ILC cell subsets 1 month post dose 2 AZD1222 (left) and 1 month post-mRNA booster (right), in individual individuals, in different age groups. Significance testing using Kruskal-Wallis one-way test. (E) T cell receptor (TCR) network of expanded T cell clones isolated from PBMCs of vaccinated individuals, by clone size, cell subset, age group and time point. (F) Proportions of CD4^+^ T cells and expanded CD4^+^ T cell clones in <70 and 70+ individuals.

Analysis of CD4 T cell scTCR data revealed several expanded TCR clones, which were enriched among *GZMA/B*-expressing cytotoxic CD4 T cells (**Figure 4E, Supplementary Figure 8**). In elderly subjects, these cytotoxic CD4 T cells comprised a greater proportion of CD4 T cells as well as expanded CD4 T cell clones than in younger individuals (**Fig. 4F**). Of note, expansion of a cytotoxic CD4+ T cell subset has been associated with increased disease severity following SARS-CoV-2 infection, but may also contribute to viral clearance ^32^.

Analysis of geneset expression showed marked differences in expression across CD4 T cell subsets between samples taken at 1 month following dose 2 AZD1222 (D2) compared with the same timepoint post-booster mRNA vaccine (D3) (**Figure 5A**). The expression of several relevant genesets, for example, ‘*interferon alpha response’*, ‘*interferon gamma response’* and ‘*IL2-STAT5 signalling’* genes was greater post-mRNA vaccine in all CD4 T cell subsets (**Fig. 5A-B**). Indeed, among TE CD4 T cells, little expression of these genesets was detectable following dose 2 AZD1222 in either age group, however, post-mRNA vaccine, there was a marked induction of ‘*IL2-STAT5 signalling’* and ‘*T cell receptor signalling’* genes, particularly in the ≥70 year age group, including *CD44* and *CD69* (**Fig. 5A-B**), consistent with our previous work on vaccine-specific TE CD4 responses in older people ^33^. In the cytotoxic CD4 T cells, the ≥70 age group showed muted expression of ‘*interferon alpha response’*, ‘*interferon gamma response’* genesets post-dose 2 AZD1222 compared with the <70 year old group. However, following the mRNA booster both age groups showed a similar enrichment of these genes (**Figure 5A-B**).

**Figure 5:**
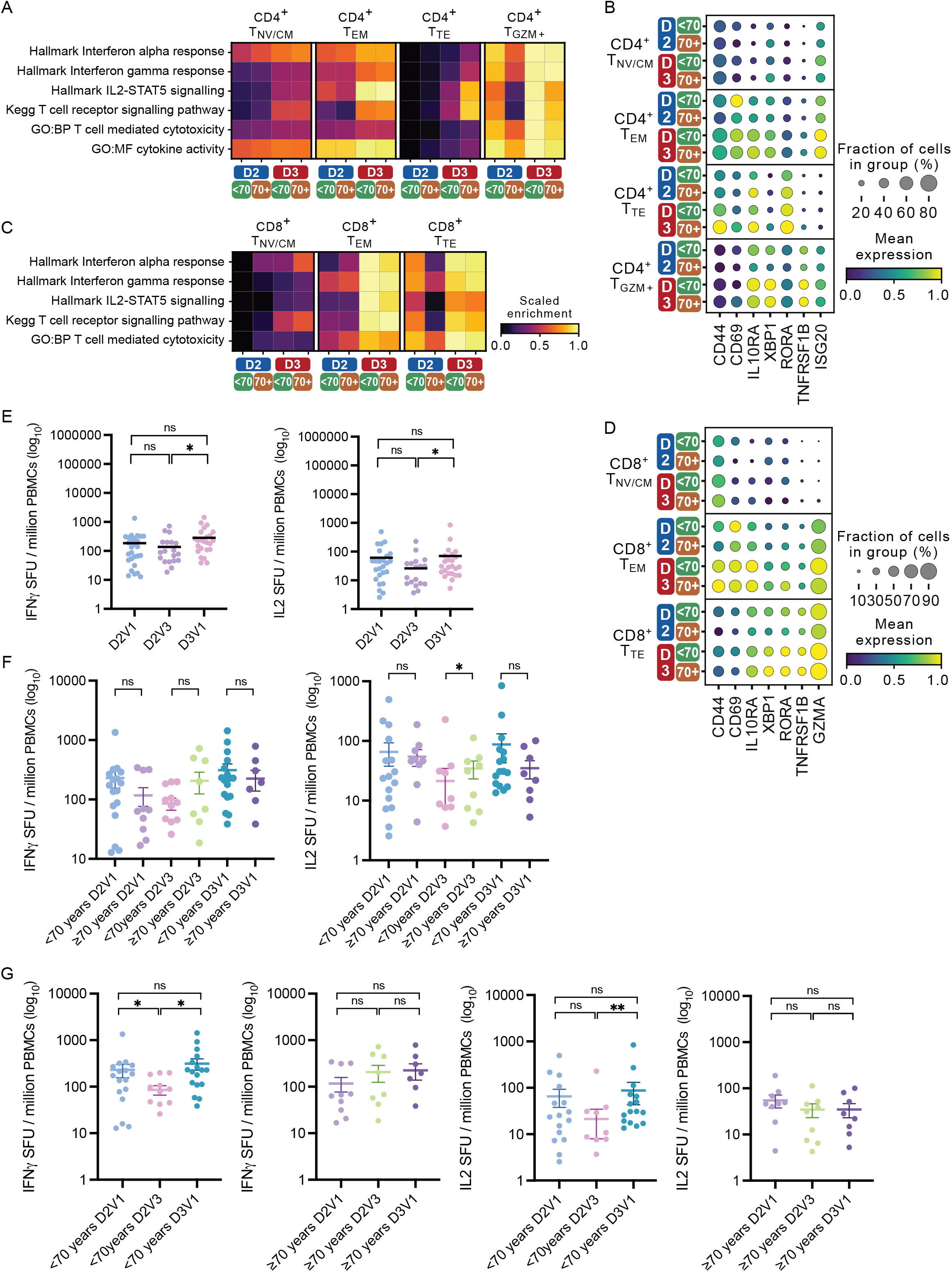
Age-associated changes in circulating T cells following AZD1222 and an mRNA booster. (A) Heatmap showing gene set expression in CD4^+^ T cell subsets in <70 and 70+ individuals 1 month post dose 2 AZD1222 (D2) and 1 month post-mRNA booster (D3). (B) Selected differentially expressed genes in CD4^+^ T cell subsets between <70 and 70+ individuals or post-D2 and D3. (C) Heatmap showing gene set expression in CD8^+^ T cell subsets in <70 and 70+ individuals 1 month post dose 2 AZD1222 (D2) and 1 month post-mRNA booster (D3). (D) Selected differentially expressed genes in CD8^+^ T cell subsets between <70 and 70+ individuals or post-D2 and D3. (E) Fluorospot analysis of IFNy and IL-2 T cell responses to SARS-CoV-2 Wu-1 D614G WT at each longitudinal time point. Wilcoxon matched-pairs signed-rank test used. NS is non-significant. * p < 0.05. (F) IFNg and IL2 SFUs per million PBMCs across longitudinal timepoints stratified by those below age 70 and those aged 70 and older. SFU: spot forming units measured by fluorospot assay. Significance testing using Mann-Whitney test (E-F). NS is non-significant, * p < 0.05. (G) IFNg and IL2 SFUs per million PBMCs by age group. Wilcoxon matched-pairs signed-rank test was used. NS is non-significant, * p < 0.05, ** p < 0.01.

In CD8 T cell subsets, several genesets showed greater expression 1 month post-mRNA booster compared with 1 month following dose 2 AZD1222 (D2) compared with the same timepoint post-booster mRNA vaccine (D3) (**Figure 5C**). In addition, in TE CD8 T cells in particular, the more muted expression observed in the ≥70 group post-D2 was reversed by the mRNA vaccine, with equivalent or even greater expression observed in the ≥70 group relative to the <70 year old group, including *GZMA* (**Figure 5C-D**).

To explore differences in antigen-specific T cell activation following the different vaccination doses and between age groups, we measured interferon gamma (IFNy) and interleukin 2 (IL-2) T cell responses in PBMCs using a Fluorospot assay. PBMCs were stimulated with overlapping peptide pools derived from the D614G SARS-CoV-2 spike and the IFNy and IL-2 responses measured. There was a significant increase in IFNn and IL2 responses following the mRNA booster compared to six months post-second dose AZD1222 (p < 0.0281 and p < 0.0291 for IFNy and IL2 respectively, **Figure 5E**). However, this difference was driven by a robust increase in T cell responses in the < 70 year age group, whilst in the ≥70 age group, no booster dose-associated augmentation in IFNy and IL2 T cell responses was evident (**Figure 5F-G, Supplementary Figure 9**). *Il2* transcripts were typically below the limit of detection in our scRNAseq data, but among CD4 T cells, some expression of *IFNG* was observed, which was greater in cells from the <70 year age group than in the ≥70 age group, as was expression of *IFNGR1* in both CD4 and CD8 T cells (**Supplementary Figure 8**).

These data indicate that T cell immunity conferred by AZD1222 persists and boosting with an mRNA-based vaccine enhances responses. However, the impact of the booster, particularly for IL-2 responses, is diminished in the elderly.

### Transcriptional changes in NK cells and myeloid cells evident after mRNA booster

Finally, we interrogated the single cell transcriptomes of the NK cells captured in our scRNAseq dataset. Circulating NK cells are comprised of two major subsets, a CD16+ subset with marked cytotoxic capacity and a CD56^bright^,CD16-subset associated with reduced cytotoxicity and prominent cytokine production, particularly Th1 cytokines such as IFNn^34^. In the CD16+ NK cells, the expression of cytotoxicity-associated genes, including *GZMB* and *PRF1* was higher at 1 month post-mRNA vaccine booster (D3) compared with 1 month post dose 2 of AZD1222 **(Supplementary Fig. 10A-B**). *FCGR3A* expression, (encoding CD16, the IgG receptor required for NK cell antibody-dependent cellular cytotoxicity (ADCC), was also higher post-D3, particularly in the ≥70 cohort (**Supplementary Fig. 10B**), potentially augmenting the anti-viral effects of the antibodies generated in the cohort. In the CD56+CD16-NK cell subset, ‘*interferon alpha response’* and ‘*interferon gamma response’* genesets were more highly expressed post-D3 compared with post-D2, but at both timepoints, expression was greater in the <70 compared with the ≥70 year old group (**Supplementary Fig. 10A**).

When considering the myeloid cells in isolation, CD14+ classical monocytes and CD16+ non-classical monocytes were the major subsets represented, with CD1c+ cDCs and pDCs the next largest populations (**Supplementary Fig. 10C-D**). The proportional representation of CD14+ monocytes decreased with age, with a corresponding increase in CD16+ monocytes with age (**Supplementary Figure 10E**), in line with previous descriptions ^35^. The activating effect of the mRNA booster on this subset was particularly remarkable in the ≥70 age group, which showed greater expression of ‘*interferon alpha response’*, ‘*interferon gamma response’*, ‘*antigen processing and presentation’* and ‘*lymphocyte co-stimulation’* genesets than that observed in the <70 years cohort (**Supplementary Fig. 10F**). cDCs, showed higher expression of ‘*antigen processing and presentation’* and ‘*lymphocyte co-stimulation’* genesets post mRNA booster (D3) compared to post-dose 2 AZD1222 (D2), the latter particularly marked CD1c+ DCs, including *CD86* and *TNFSF13B* (encoding BAFF) (**Supplementary Fig. 10G-H**). In pDCs, there was also higher expression of ‘*interferon alpha and beta production’* genesets post-D3 compared with post-D2 (**Supplementary Fig. 10I**).

A previous analysis of responses to second dose of mRNA vaccine (following primary mRNA vaccine dose) found that early monocyte activation correlated with the development of SARS-CoV-2 neutralizing antibodies and CD8 T cell IFNn responses ^36^. Altogether, our analysis suggests that even a month after the booster mRNA vaccine, there is evidence of on-going transcriptional activation of monocytes, pDCs and cDCs, with expression of several genes that may promote T and B cell activation. In contrast to adaptive immune cells, myeloid cells do not exhibit classical immunological memory. Therefore, the enhanced myeloid cell activation observed in response to the mRNA vaccine relative to dose 2 AZD1222 likely reflects a vaccine-intrinsic feature.

## Discussion

Long term vaccine elicited immunity is important for protection against SARS-CoV-2 variants, and can be measured by circulating binding and neutralizing antibodies, spike specific T cell immunity, and spike specific B cell responses ^15,37^. Neutralizing antibody levels wane over time, with a significant decrease seen six months post second dose ^38,39^. In contrast, T cell immunity is longer lived and may confer durable protection, even as new variants emerge. Studies showed that the T cell response remained robust over a six-month period, even to Omicron BA.1 ^18,39,40^.

Compared to mRNA primary course vaccination, two dose AZD1222 vaccine has been shown to confer poorer protection against infection with variants of concern including Beta ^41^, and Delta, with breakthrough cases emerging ^6,23,42^ even when peak antibody titres are expected. With titres of neutralizing antibodies waning in the general population after mRNA or adenovirus vectored vaccine primary course ^43,44^, an mRNA booster was recommended based on early studies with mRNA as the third vaccine dose; previous studies ^22,45-48^ showed that heterologous vaccination in individuals primed with AZD1222, AD26.COV2.S and boosted with an mRNA-based vaccine, or homologous vaccination with BNT162b2, enhanced immune responses as determined by measurement of neutralising antibodies and T cell responses. Additionally, the booster vaccine dose aided seroconversion in immunosuppressed individuals ^49^. However, little *in vitro* data exists on boosting in the elderly population, in contrast to epidemiological data ^22,50,51^; this lack of data is particularly evident for heterologous prime-boost approaches ^45^.

Primary course AZD1222 vaccine was used after BNT162b2 in the UK and therefore in younger individuals between the ages of 40 and 75. Following early data on boosting of immune responses after mRNA third dose ^22,47^ mRNA-based vaccines were offered as a booster vaccine 6 months after primary two dose courses of either AZD1222 or BNT162b2. In our cohort of 36 individuals, 13 whom were 70 or older, we assessed binding and neutralizing responses as well as T cell and B cell responses to vaccination over time. Significant waning of neutralizing antibodies was observed across all individuals 6 months post second-dose, but 1 month after mRNA-based booster vaccination, titres increased significantly, to levels that were also significantly higher than those seen 1 month after the second dose of AZD1222. Interestingly, no differences were observed between age groups for dose 1 and 2. However, following booster vaccination, the ≥70 year old group did not respond as well as the under 70 group. However, whilst age-related differences were observed in the neutralization, total Spike IgG levels showed no association with age. This pointed towards differences in neutralization potency and possibly breadth, rather than quantity of spike specific antibody. We also observed suboptimal boosting of spike specific T cell responses in the elderly after dose 3 that was most marked for the IL2 response.

Phenotyping RBD-specific B cells from one-month post-boost revealed a distinct population of IgD– RBD+ age-associated atypical memory B cells that was present at a higher frequency in older individuals than younger participants. The literature surrounding atypical memory B cells describes various roles in humans, although these different functions may be context-dependent ^52^. Initially, B cells with this phenotype were characterized as exhausted or hypo-responsive memory B cells that formed after infection or in autoimmune disease^53-55^. Additionally, there an accumulation of atypical memory B cells in older individuals, suggesting that biological changes that occur with age can favor skewing of the memory B cell pool towards atypical B cells ^26,56^. The formation of atypical memory B cells can be supported by IL-21 and IFNy, and be inhibited by IL-4 ^26^, therefore, these cells may emerge as a natural consequence of the increased inflammation that is present in older people. We have previously shown that hemagglutinin-specific Tfh cells that are induced by vaccination have an enhanced IFNγ gene signature in older donors ^33^, indicating that atypical B cell promoting conditions exist in older people upon vaccination.

Although first described in immune pathology, it is now clear that atypical memory B cells emerge from normal B cell activation in response to vaccination^52,56-59^. Most studies suggest that the majority of atypical B cells are non-GC derived ^56,60,61^. We have previously described that the AZD1222 elicits a diminished GC response in aged mice compared to younger animals ^62^. The poor GC observed in older individuals may necessitate the emergence of memory B cells via the extra-follicular pathway, which could explain the increased frequency of atypical memory B cells in the elderly observed in this study ^56^. This pathway may not be as favoured in younger individuals due to their more functional GC responses, reliable memory B cell populations, and resultant greater titres of neutralizing antibodies from earlier immunisations. Nonetheless, whether poor immunity results from an accrual of atypical memory B cells, or whether atypical memory B cells are generated due to a waning immune system, this increased frequency in older individuals reflects a clear difference in the immune response one month following a booster dose. It remains to be seen whether this dysfunctional state can be pharmacologically addressed, or overcome by increased dosing.

Limitations of our study include relatively modest sample size, sampling of peripheral blood to measure vaccine induced immune responses and lack of clinical data on protection from subsequent COVID-19 and severity.Going forward, it will be important to understand the dynamics of waning in elderly individuals, as well as the impact of the fourth dose and differences by age. Such studies are increasingly challenging due to the heterogeneity in time intervals between vaccine doses. Nonetheless, the elderly remain a key target population for maximizing protective vaccine responses as they are still disproportionally likely to have poor health outcomes after SARS-CoV-2 infection, warranting continued comprehensive assessment.

## Data Availability

All data produced in the present study are available upon reasonable request to the authors

## Acknowledgements

We would like to thank Cambridge University Hospitals NHS Trust Occupational Health Department. We would also like to thank the NIHR Cambridge Clinical Research Facility and staff at CUH, the Cambridge NIHR BRC Stratified Medicine Core Laboratory NGS Hub, Petra Mlcochova, Martin Potts, Ben Krishna, Marianne Perera and Georgina Okecha. We thank Dr James Voss for the kind gift of HeLa cells stably expressing ACE2. RKG is supported by a Wellcome Trust Senior Fellowship in Clinical Science (WT108082AIA). This research was supported by the National Institute for Health Research (NIHR) Cambridge Biomedical Research Centre, the Cambridge Clinical Trials Unit (CCTU), the NIHR BioResource and Addenbrooke’s Charitable Trust, the Evelyn Trust (20/75), UKRI COVID Immunology Consortium. This study was supported by Biotechnology and Biological Sciences Research Council funding to MAL (BBS/E/B/000C0427, BBS/E/B/000C0428), and the Campus Capability Core Grant to the Babraham Institute.The views expressed are those of the authors and not necessarily those of the NIHR or the Department of Health and Social Care. IATMF is funded by a SANTHE award. MAL is an EMBO Young Investigator and Lister Institute Prize Fellow.

## Methods

### Study Design

Study participants were recruited from the community after receiving their second dose of the AZD1222 vaccine between the 7^th^ of April and the 22^nd^ of May 2021. They were recruited at Addenbrookes Hospital into the COVID-19 cohort of the NIHR Bioresource. Participants were followed up 1 month after receiving their second dose of the AZD1222 vaccine to provide blood samples. Participants were followed up again for a blood sample at 6 months post second dose. Once mRNA booster vaccines were made available to the community, participants were followed up for a further blood sample one month after the booster. The group of participants were categorized into two study groups as age was of interest: those <70 and those ≥70 years. The outcome of interest was vaccine-elicited serum antibody neutralisation activity at each time point and the effect of longitudinal waning and boosting on this activity. This was measured as the dilution of serum required to inhibit infection by 50% (ID50) in an *in vitro* neutralization assay. An ID50 of 20 or below was used as a cut-off to determine insufficient neutralisation. The binding antibody response to Spike protein were measured by multiplex particle-based flow cytometry. T cell responses were measured by IFNγ and IL-2 FLUOROSPOT assays.

### Ethical approval

The study was approved by the East of England – Cambridge Central Research Ethics Committee (17/EE/0025). PBMC from unexposed volunteers previously recruited by the NIHR BioResource Centre Cambridge through the ARIA study (2014-2016), with ethical approval from the Cambridge Human Biology Research Ethics Committee (HBREC.2014.07) and currently North of Scotland Research Ethics Committee 1 (NS/17/0110).

### Statistical Analyses

Descriptive analyses of demographic and clinical data are presented as median and interquartile range (IQR) when continuous. When categorical, these data are presented as frequency and proportion (%). Linear regression was used to model the association between age and S total IgG at each time point as well as the association between S total IgG and ID50 for the same time point. Pearson’s correlation was used to measure the relationship between the variables. Linear regression was also used to measure the association between IFNy and ID50. Statistical analyses were run using GraphPad Prism. UMAP analysis was performed using R (version 4.1.1) using code that has previously been described ^63^.

### Generation of Mutants and pseudotyped viruses

Wild-type (WT) bearing 614G, B.1.617.2, and B.1.1.529 pseudotyped viruses were generated as previously described ^24^. In brief amino acid substitutions were introduced into the D614G pCNA_SARS-CoV-2_S plasmids as previously described ^64^. The pseudoviruses were generated in a triple plasmid transfection system whereby the Spike expressing plasmid along with a lentviral packaging vector-p8.9 and luciferase expression vector-psCSFLW where transfected into 293T cells with Fugene HD transfection reagent (Promega). The viruses were harvested after 48 hours and stored at -80°C. TCID50 was determined by titration of the viruses on 293Ts expressing ACE-2 and TMPRSS2.

### Neutralisation assays

Virus neutralisation assays were run using HeLa expressing ACE2 cells using SARS-CoV-2 Spike pseudotyped virus expressing luciferase. Pseudotyped virus was incubated with serially diluted heat inactivated human serum samples or sera from vaccinees in duplicate for 1h at 37°C. Cell only and virus and cell only controls were included. After an hour, HeLa ACE2 cells were added to each well. Following 48h of incubation at 5% CO_2_ and 37°C, luminescence was measured using the BrightGlo^™^ Luciferase Assay System (Promega, UK). Neutralization was calculated relative to the virus and cell only controls. Data was analysed in GraphPad Prism where 50% neutralisation (ID50) values were calculated and the limit of detection for neutralisation was set at an ID50 of 20. Within each group, the ID50 values were summaraised a geometric mean titre (GMT). Statistical comparisons between groups were made using either the Wilcoxon ranked sign test or the Mann-Whitney test.

### SARS-CoV-2 serology by multiplex particle-based flow cytometry (Luminex)

Recombinant SARS-CoV-2 N, S and RBD were covalently coupled to distinct carboxylated bead sets (Luminex; Netherlands) to form a 3-plex and analyzed as previously described ^24^. Specific binding was reported as mean fluorescence intensities (MFI).

### Serum autoantibodies

Serum was screened for the presence of autoantibodies using the ProtoPlexTM autoimmune panel (Life Technologies) according to the manufacturer’s instructions. Briefly, 2.5ml of serum was incubated with Luminex MagPlex magnetic microspheres in a multiplex format conjugated to 19 full length human autoantigens (Cardiolipin, CENP B, H2a(F2A2) & H4 (F2A1), Jo-1, La/SS-B, Mi-2b, myeloperoxidase, proteinase-3, pyruvate dehydrogenase, RNP complex, Ro52/SS-A, Scl-34, Scl-70, Smith antigen, Thyroglobulin, Thyroid peroxidase, transglutaminase, U1-snRNP 68, whole histone) along with bovine serum albumin (BSA). Detection was undertaken using goat-anti-human IgG-RPE in a 96 well flat-bottomed plate and the plate was read in a Luminex xMAP 200 system. Raw fluorescence intensities (FI) were further processed in R (version 3.5.1) Non-specific BSA-bound FI was subtracted from background-corrected total FI for each antigen before log_2_ transformation and thresholding. Outlier values (Q3+1.5*IQR) in each distribution were defined as positive.

### IFNγ and IL2 FLUOROSPOT T cell assays

Peripheral blood mononuclear cells (PBMC) were isolated from the heparinized blood samples using Histopaque-1077 (Sigma-Aldrich) and SepMate-50 tubes (Stemcell Technologies). Frozen PBMCs were rapidly thawed and diluted into 10ml of TexMACS media (Miltenyi Biotech), centrifuged and resuspended in 10ml of fresh media with 10U/ml DNase (Benzonase, Merck-Millipore via Sigma-Aldrich), PBMCs were then incubated at 37°C for 1h, followed by centrifugation and resuspension in fresh media supplemented with 5% Human AB serum (Sigma Aldrich) before being counted. PBMCs were stained with 2ul of LIVE/DEAD Fixable Far Red Dead Cell Stain Kit (Thermo Fisher Scientific) and live PBMC enumerated on the BD Accuri C6 flow cytometer.

1.0 to 2.5 × 10^5^ PBMCs were incubated in pre-coated FluoroSpot^FLEX^ plates (anti IFN*γ*and IL2 capture antibodies Mabtech AB, Nacka Strand, Sweden)) in duplicate with either peptide mixes specific for Wuhan-1(QHD43416.1) Spike SARS-CoV-2 protein (Miltenyi Biotech) or a mixture of peptides specific for Cytomegalovirus, Epstein Barr virus and Influenza virus (CEF+) (final peptide concentration 1μg/ml/peptide, Miltenyi Biotech) in addition to an unstimulated (media only) and positive control mix (containing anti-CD3 (Mabtech AB) and Staphylococcus Enterotoxin B (SEB), (Sigma Aldrich) at 37ºC in a humidified CO2 atmosphere for 42 hours. The cells and medium were then decanted from the plate and the assay developed following the manufacturer’s instructions. Developed plates were read using an AID iSpot reader (Oxford Biosystems, Oxford, UK) and counted using AID EliSpot v7 software (Autoimmun Diagnostika GmbH, Strasberg, Germany). Peptide specific frequencies were calculated by subtracting for background cytokine specific spots (unstimulated control) and expressed as SFU/Million PBMC.

### Sample processing, library preparation, and sequencing

PBMC samples were removed from -80 storage and defrosted by gradual addition and removal of ice-cold PBS, resuspending the frozen cells to a final volume of 40 mL while keeping the samples on wet ice throughout defrosting. The cells were centrifuged at 400g for 5 minutes. The supernatant was discarded, and cells were re-suspended in a small volume of PBS with CaCl_2_, as required for enrichment of live cells, using EasySep (STEMCELL technologies) dead cell removal kit, following the manufacturer’s instructions. Following this, cells were centrifuged as before and counted. Two or three samples from distinct individuals were pooled (i.e. genotype multiplexing) in an overlapping mixture design at equal concentrations, counted, and 1×10^5^ cells were resuspended in 100μL of PBS.

### The 10x Chromium GEM Single Cell V(D)J 5’ kit v2 (dual index) with BCR and TCR

amplification was used for library preparation. Samples were loaded onto the chip following the manufacturer’s recommendations, with an aim to recover 8000 cells (for 2 samples) or 12000 cells (for 3 samples) per lane. The remainder of the 10x library preparation was carried out as per manufacturer’s instructions and the resulting libraries (GEX, TCR, BCR) sequenced using NovaSeq 6000 paired-end sequencing (Illumina) at Genewiz. BCL files were demultiplexed using Casava (Illumina) and count tables produced using CellRanger v7.0 (10x genomics).

### Single-cell RNAseq data and pre-processing

Genotype demultiplexing was performed using Souporcell (v2) ^65^. Souporcell analyses was performed using the ‘skip_remap’ setting and a set of known donor genotypes given under the ‘common_variants’ parameter, and the k number set at the number of samples loaded per lane. The donor ID for each Souporcell genotype cluster was annotated by comparing with known genotypes from the multiplex design, using GenotypeMixtures ^6666^Droplets containing more than one genotype according to Souporcell or with unresolved genotypes were removed. Further doublet detection was performed on the combined raw count data (10x CellRanger output) using Scrublet (v0.2.3) ^67^. Following this, iterative sub-clustering was performed, the median Scrublet score for each sub-cluster was computed, and median absolute deviation scores were calculated followed by application of a one-tailed t-test with Benjamin-Hochberg correction, as previously described ^68^. Cells with significantly outlier Scublet scores (corrected Pval < 0.1) were regarded as probable doublets and filtered. The data was then processed using Scanpy following the standard workflow ^69^. Cells were filtered if they contained >200 or <8000 genes. Percentage mitochondrial content cut-off was set at <15%. Genes were retained if they were expressed in three or more cells. Highly variable genes were selected based on a minimum and maximum expression of >0.0125 and <3 respectively; with the minimum dispersion of genes = 0.5. TCR and BCR V(D)J genes were removed from highly variable genes. The number of PCs used for neighbourhood graph construction and dimension reduction was set at 30. Batch correction was performed using bbknn using the ridge regression setting and 10x sequencing lane as the batch term ^70,71^. Clustering was performed using the Leiden algorithm ^72^. Visualisation of reduced dimensions was performed with UMAP (v3.10.0) using a minimum distance of 0.3 and all other parameters according to the default settings in Scanpy ^73^ For initial clustering, differentially expressed genes were calculated using the Wilcoxon rank-sum test. Finally, cell clusters expressing improbable combination of cell type markers were filtered, after manual inspection of the data. This led to a working data set of 99,384 cells.

### Single-cell gene expression analysis

Preliminary annotation of cell clusters was performed with CellTypist ^25^. Briefly, the ‘Covid19 immune landscape’ model was used to predict cell-types based on logistic regression classifiers, using the majority voting classifier setting. Next, clusters were manually inspected, to obtain the final annotations using a combination of canonical mRNA markers and BCR/TCR sequencing information, where available. Gaussian kernel density estimation was performed using Scanpy’s tl.embedding_density function. Compositional analysis was performed using scCODA, which applies a Bayesian model to identify cell type changes ^74^. Gene sets were obtained from the Molecular Signature Database (MSigDB v7.3) inventory ^75^. Gene signature scoring was performed with UCell, which is based on the Mann-Whitney U statistic ^76^. For patient-level comparisons, cell-level scores were averaged (mean) by sample, for each cell type. Mann-Whitney U test was applied for age comparisons or Wilcoxon signed-rank test for dose comparisons, where paired patient samples were available.

### BCR and TCRseq analysis

Single-cell V(D)J data was processed with the CellRanger vdj v7.0 pipeline. BCR and TCR V(D)J data was processed using dandelion (v0.2.4), as previously described ^77^. Briefly, individual BCR contigs were reannotated using the IMGT reference database ^78^, and heavy-chain V-gene alleles corrected for individual genotypes using TIgGER ^79^. Constant genes were re-annotated using blastn (v2.10.0) with CH1 regions from IMGT. For BCR filtering, the ‘ddl.pp.check_contigs’ function was used to remove poor quality or ambiguous contigs from the V(D)J data but the transcriptome barcode was retained. Clones were defined as previously described, and the resulting data was combined with gene expression for further analysis.

For identification of putative BCR sequences with capacity to bind SARS-CoV2 surface antigens, we downloaded CDR3 sequences and germline assignments of known SARS-CoV2-binding antibodies from the Coronavirus Antibody Database (CoVAbDab, updated 26/07/22), and filtered for antibodies of B cell origin ^28^. CDR3 sequences assessed because CDR3 plays a dominant role in antigen binding and specificity ^80,81^. We defined putative SARS-CoV2-binding BCR sequences in our data set as B cells with complete CDR3 light-chain (CDR3-L) match and significant motif similarity for CDR3 heavy-chain (CDR3-H) compared to the CoVAbDab database. Motif enrichment analysis between BCR CDR3-H sequences in our data and CoVAbDab was performed using gliph2 ^82^. First, we compiled a reference database of naïve BCR CDR3 sequences using published CDR3 sequence from Covid-naïve B cells from multiple sources, for the gliph2 pipeline. Briefly, 1,437,743 CDR3-H sequences from bulk BCR-sequencing (downloaded from iReceptor gateway 30/07/22) ^83^ and 55,444 CDR3-H sequences from three single-cell GEX + V(D)J-seq studies ^77,84,85^ of control Covid-naïve B cells were included in the reference. For gliph2 analysis, we considered only motifs that were over 5 residues in length. We assessed motifs biased towards the target data (CoVAbDab + BCRseq V(D)J data combined) versus the naïve reference data (Fisher exact test p < 0.05), to identify sequences enriched post-vaccination in our cohort. Shared motifs were visualised using ggseqlogo ^86^. Our approach gave us 1460 single-cell CDR3-L sequences identical to and 434 single-cell CDR3-H with shared motifs to SARS-CoV2-binding CDR3-H/L sequences from CoVAbDab. We recombined CDR3-H and CDR3-L sequences using their single-cell barcodes and filtered for single cells where both CDR3-H+L sequences matched the corresponding CoVAbDab antibody. This led to 152 ‘double-matched’ putative SARS-CoV2-binding single-cell BCRs.

## Supplementary Figures

**Supplementary figure 1: Table of characteristics of study participants and neutralization data for Wu-1 D614G WT**.

**Supplementary figure 2: S antibody responses elicited by AZD122 vaccine and mRNA booster**. (A) Linked S antibody responses from longitudinal time points across individuals. (B) S antibody responses in the <70 population and the >70 population. Wilcoxon matched-pairs signed ranked test was used. (C) Linked S antibody responses in the <70 population and the >70 population. Wilcoxon matched-pairs signed ranked test was used.

**Supplementary figure 3: Longitudinal neutralizing plasma antibody titers against Wu-1 D614G WT, Delta, and Omicron variants from AZD1222 vaccinated individuals boosted with an mRNA-based vaccine**. (A) Linkage of neutralising antibodies from longitudinal time points across individuals in response to different variants. (B) Neutralising antibody titers from the <70 group at each time point against WT, Delta, and Omicron. Wilcoxon matched-pairs signed ranked test was used. (C) Neutralising antibody titers from the >70 group at each time point against WT, Delta, and Omicron. Wilcoxon matched-pairs signed ranked test was used. (D) Linkage of neutralising antibodies from longitudinal time points from the <70 group against WT, Delta, and Omicron. Wilcoxon matched-pairs signed ranked test was used. (E) Linkage of neutralising antibodies from longitudinal time points from the >70 group against WT, Delta, and Omicron. Wilcoxon matched-pairs signed ranked test was used.

**Supplementary figure 4: Supporting data for full scRNAseq dataset**. (A) UMAP of all cells captured by scRNAseq post quality control and filtering, by individual genotype. (B) CellTypist annotation of scRNAseq, used for preliminary coarse cell-type identification. (C) Canonical marker gene expression from (A).

**Supplementary figure 5: Supporting data for scRNAseq of B cell subset**. (A) CellTypist annotation of atypical memory B cells. (B) Atypical memory B cells express*FCRL5* and lack *CR2*. (C) Ig heavy-chain isotype calls from matched scBCRseq.

(D) Selected differentially expressed genes driving differences in ‘Antigen processing and presentation’ in B cells, from figure 2G. (E) *IL21R* and *IFNGR1* expression in B cell subsets in <70 and 70+ individuals 1 month post dose 2 AZD1222 (D2) and 1 month post-mRNA booster (D3).

**Supplementary figure 6: BCRseq analysis**. (A) Summary strategy for identifying putative SARS-CoV-2-binding BCR sequences; and scBCRs with matching CDR3-light chain and heavy chain matches to CoV-AbDab, before combining to identify CDR3-heavy+light chain matches in figure 3A. (B) CDR3 heavy chain sequences of memory B cells from individuals post-vaccination are enriched for highly-similar motifs to CDR3 heavy chain sequences from CoV-AbDab, particularly at the central residues. (C) Proportions of SARS-CoV-2-binding B cells by memory B cell subset, in <70 and 70+ individuals.

**Supplementary figure 7: Flow cytometry gating for spike-specific B cell phenotyping**. For supplementary figure 5, a representative <70-year-old D3V1 sample was used. (A) Flow cytometry gating strategy used to define IgD-B cells. (B) Flow cytometry gating of RBD+ cells (pre-gated on IgD-B cells as in (A)). (C) Flow cytometry gating of Spike+ cells (pre-gated on IgD-B cells as in (A)). (D) Flow cytometry gating of CD11c+ FCRL5+ cells (pre-gated on B cells as in (A). (E) Flow cytometry gating of CD11c+ FCRL5+ cells (pre-gated on RBD+ IgD-B cells as in (A). (F) Flow cytometry gating of CD11c+ FCRL5+ cells (pre-gated on Spike+ IgD-B cells as in (A)

**Supplementary figure 8: Supporting data for scTCR/RNAseq of T/NK/ILC cell subset**.

(A) TCR network of expanded T cell clones isolated from PBMCs of vaccinated individuals, by genotype (corresponding to individual study participants). (B) *IL2, IL2RA, IFNG, IFNGR1* expression in CD4^+^ and CD8^+^ T cell subsets in <70 and 70+ individuals 1 month post dose 2 AZD1222 (D2) and 1 month post-mRNA booster (D3).

**Supplementary figure 9: Spike specific peripheral T cell responses**.

(A) IFNγ and IL2 SFUs per million PBMCs stratified by age of 1 month post second dose compared to one month post booster for IFNγ and IL-2 expression. (B) IFNγ and IL2 SFUs per million PBMCs stratified by age of 6 months post second dose and one month post booster for IFNγ and IL-2 expression. SFU: spot forming units measured by fluorospot assay.

**Supplementary figure 10: Age-associated changes in circulating NK and myeloid cells following AZD1222 and an mRNA booster**. (A) Heatmap showing gene set expression in NK cell subsets in <70 and 70+ individuals 1 month post dose 2 AZD1222 (D2) and 1 month post-mRNA booster (D3). (B) Selected differentially expressed genes driving differences in (A). (C) UMAP of subsetted myeloid cells annotated by canonical marker gene expression (D). (E) Proportions of myeloid cell subsets 1 month post dose 2 AZD1222 (left) and 1 month post-mRNA booster (right), in individual individuals, in different age groups. Significance testing using Kruskal-Wallis one-way test. (F) Heatmap showing gene set expression in monocytes in <70 and 70+ individuals 1 month post dose 2 AZD1222 (D2) and 1 month post-mRNA booster (D3). (G) Heatmap showing gene set expression in conventional DCs in <70 and 70+ individuals 1 month post dose 2 AZD1222 (D2) and 1 month post-mRNA booster (D3). (H) Selected differentially expressed genes driving differences in (G). (I) Heatmap showing gene set expression in conventional DCs in <70 and 70+ individuals 1 month post dose 2 AZD1222 (D2) and 1 month post-mRNA booster (D3).

