## Supplementary figures for "Atypical B cells and impaired SARS-CoV-2 neutralisation following booster vaccination in the elderly"

#### Supplementary figure 1

| | <70 | ≥70 | |
|---|---|---|---|
| **n** | 23 | 13 | |
| **Female %** | 69.6 | 69.2 | |
| **Median age (IQR)** | 66 | 73 | |
| **Sera GMT WT** | | | |
| **D2V1** | 408.4 (157 - 1063) | 266 (111.4 - 635.5) | |
| **D2V3** | 179.9 (86.11 - 376) | 97.68 (35.54 - 268.5) | |
| **D3V1** | 5749 (1683 - 19633) | 1093 (228.8 - 5218) | |
| **Sera ID50<20** | | | |
| **D2V1** | 21 (6/23) | 8 (1/13) | |
| **D2V3** | 21 (6/23) | 11 (2/13) | |
| **D3V1** | 7 (2/23) | 8 (1/13) | |
| **Time between dose 2 & dose 3** | 200.5 | 201.5 | median = 200.5 |

### Supplementary figure 2

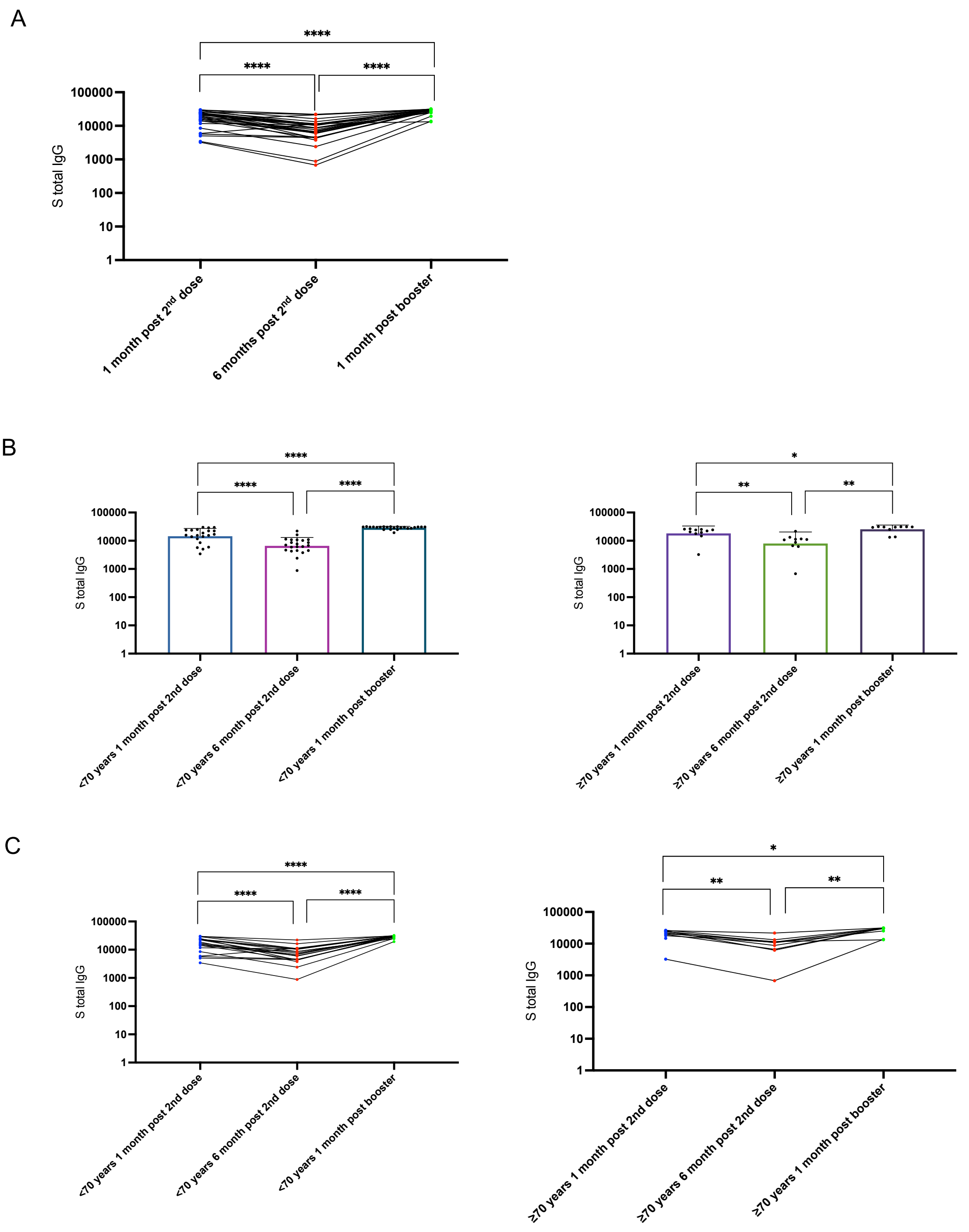

### Supplementary figure 3

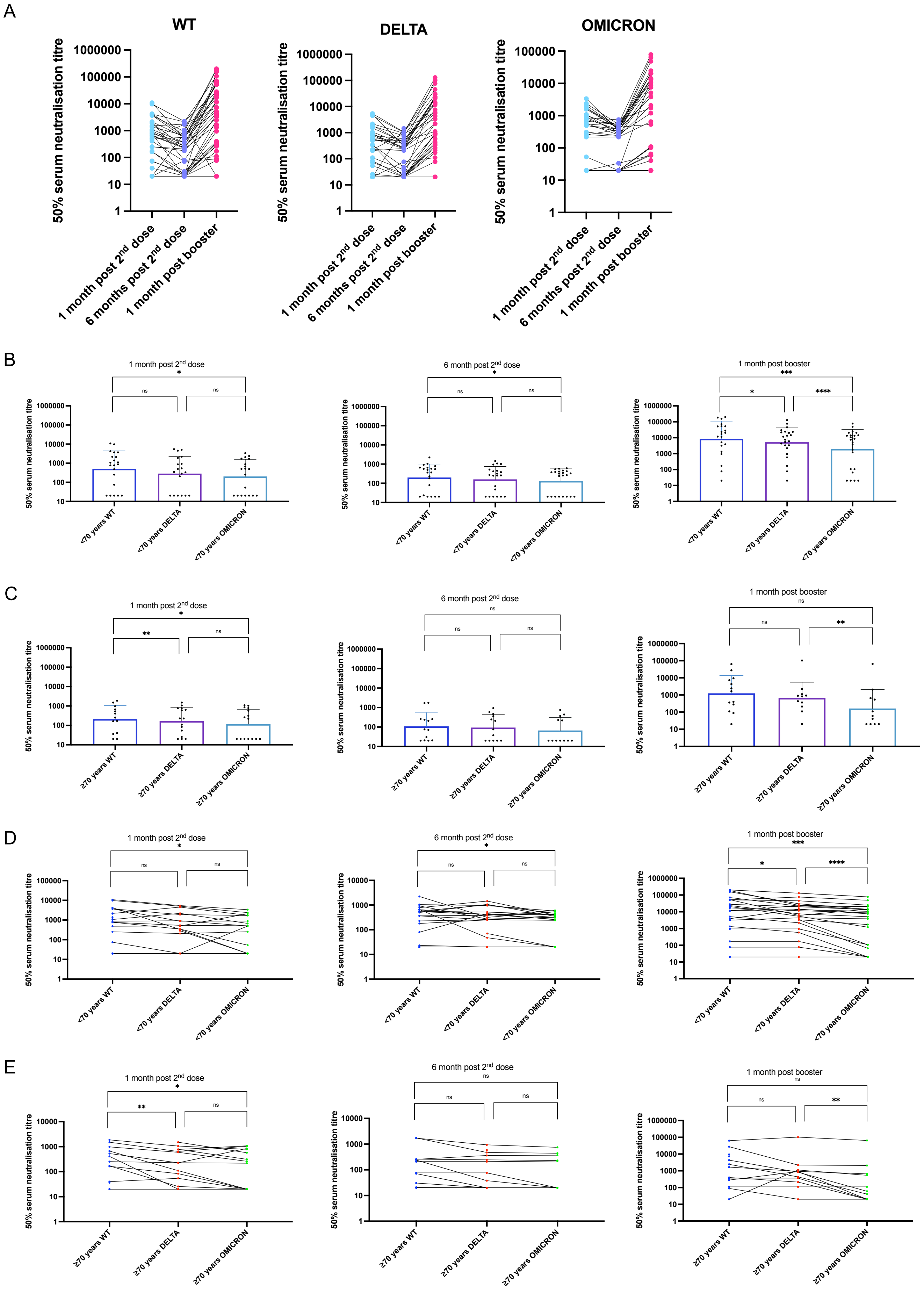

### Supplementary figure 4

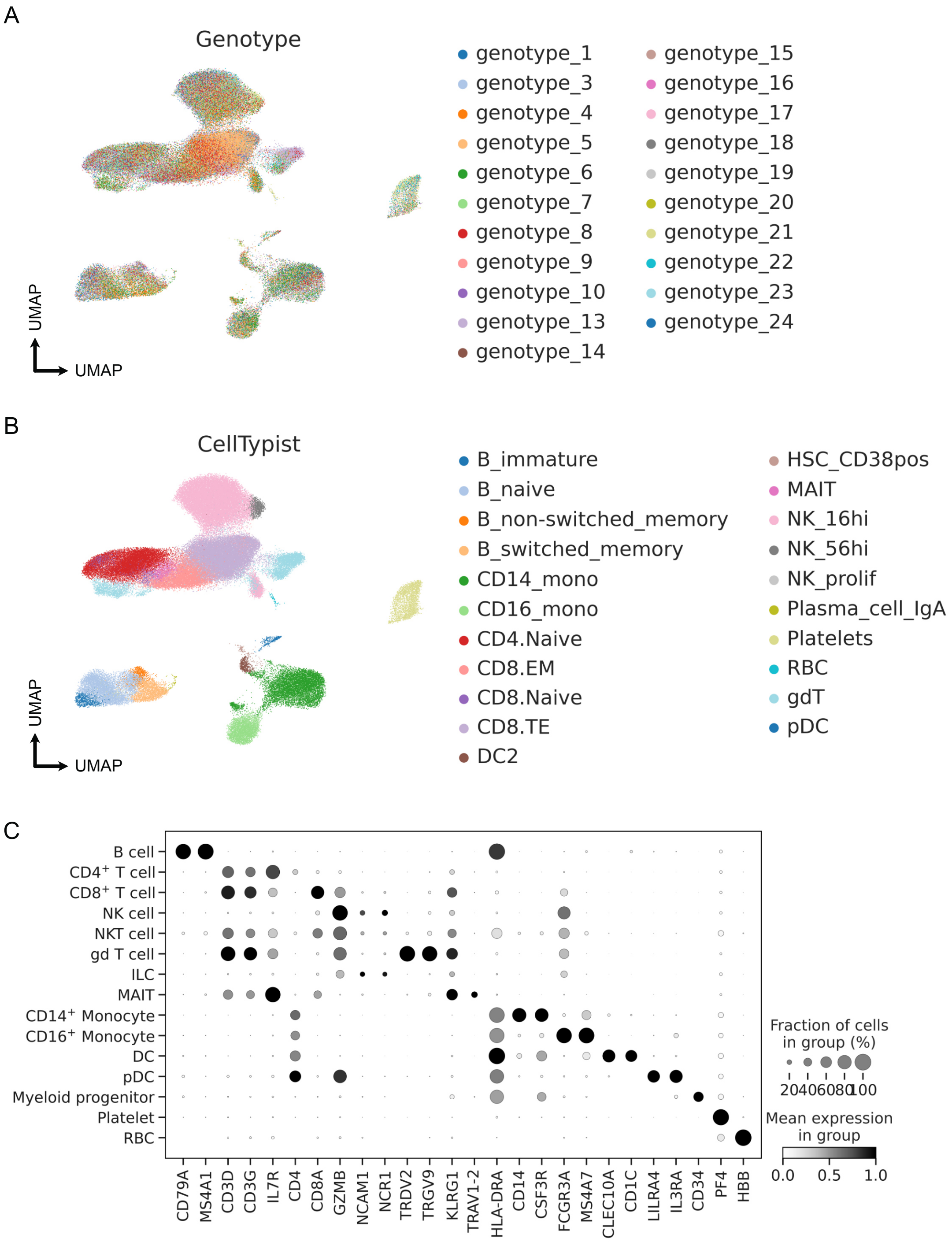

### Supplementary figure 5

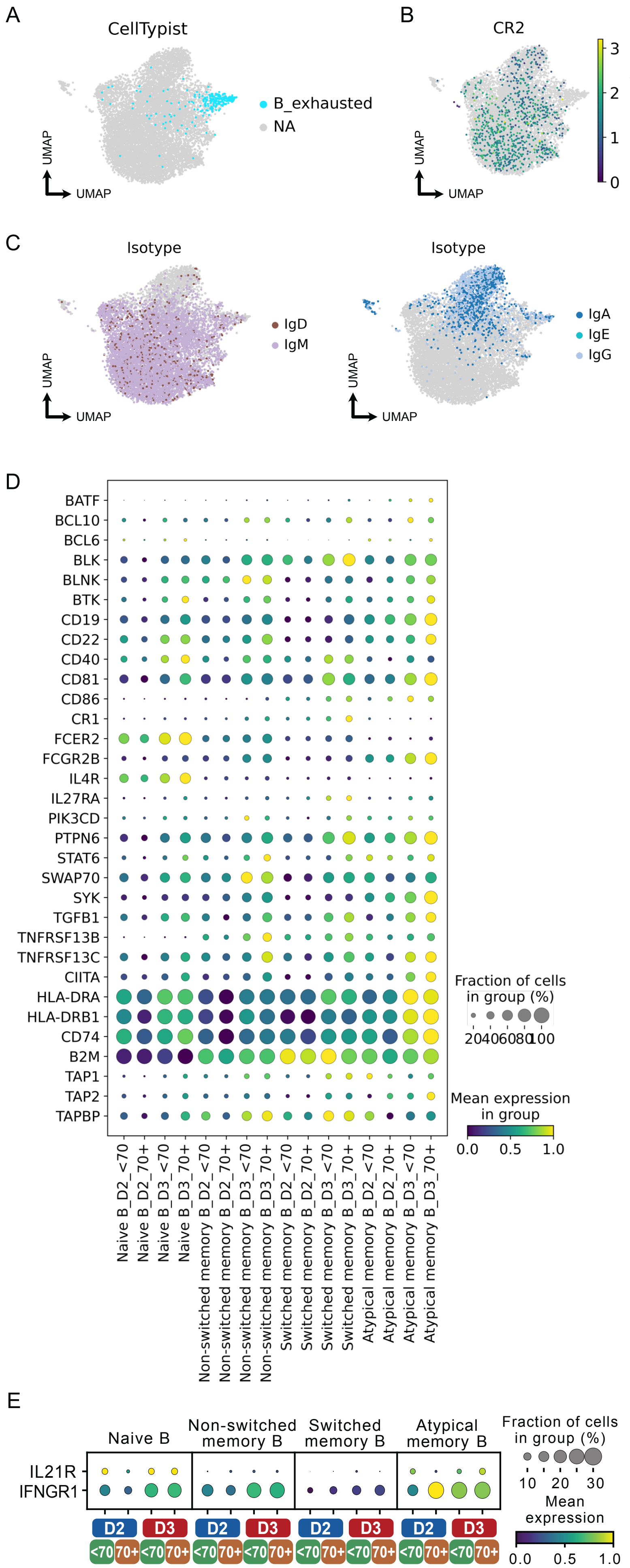

### Supplementary figure 6

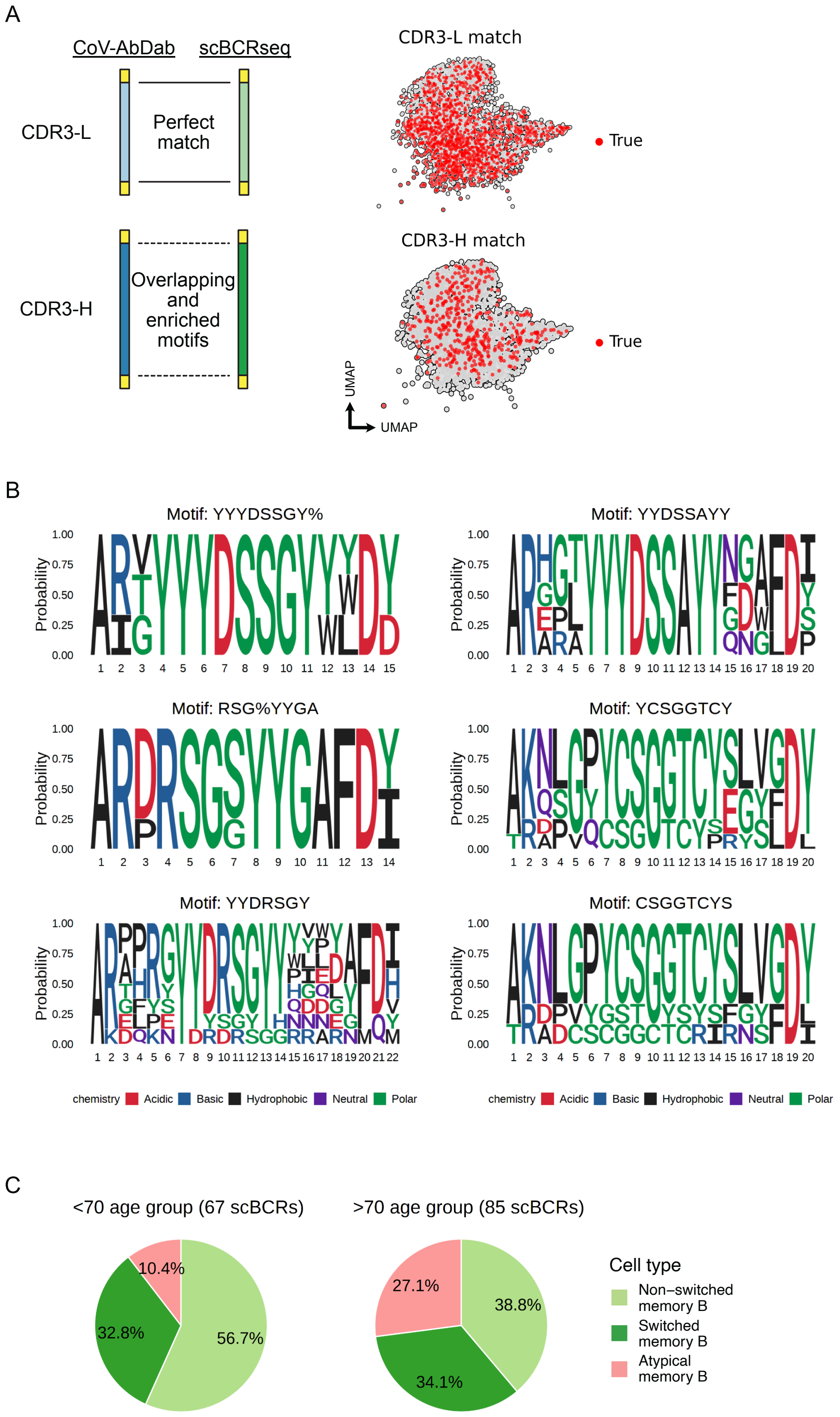

#### Supplementary figure 7

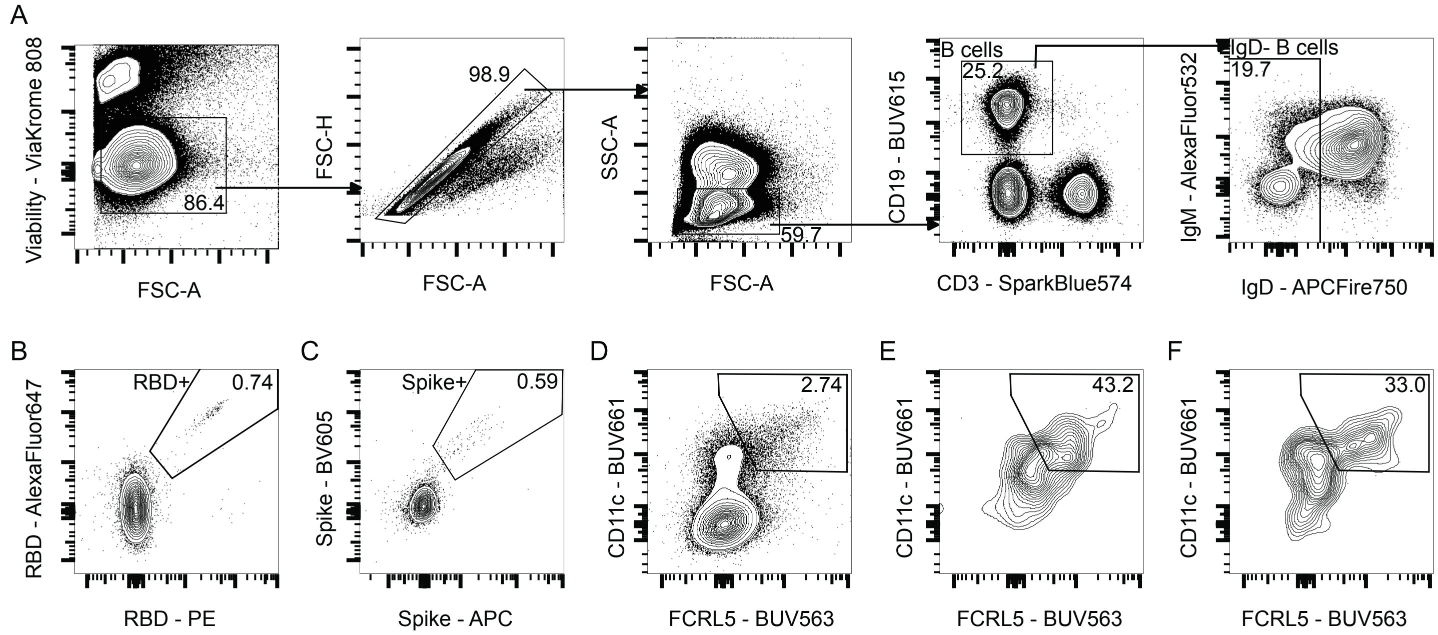

### Supplementary figure 8

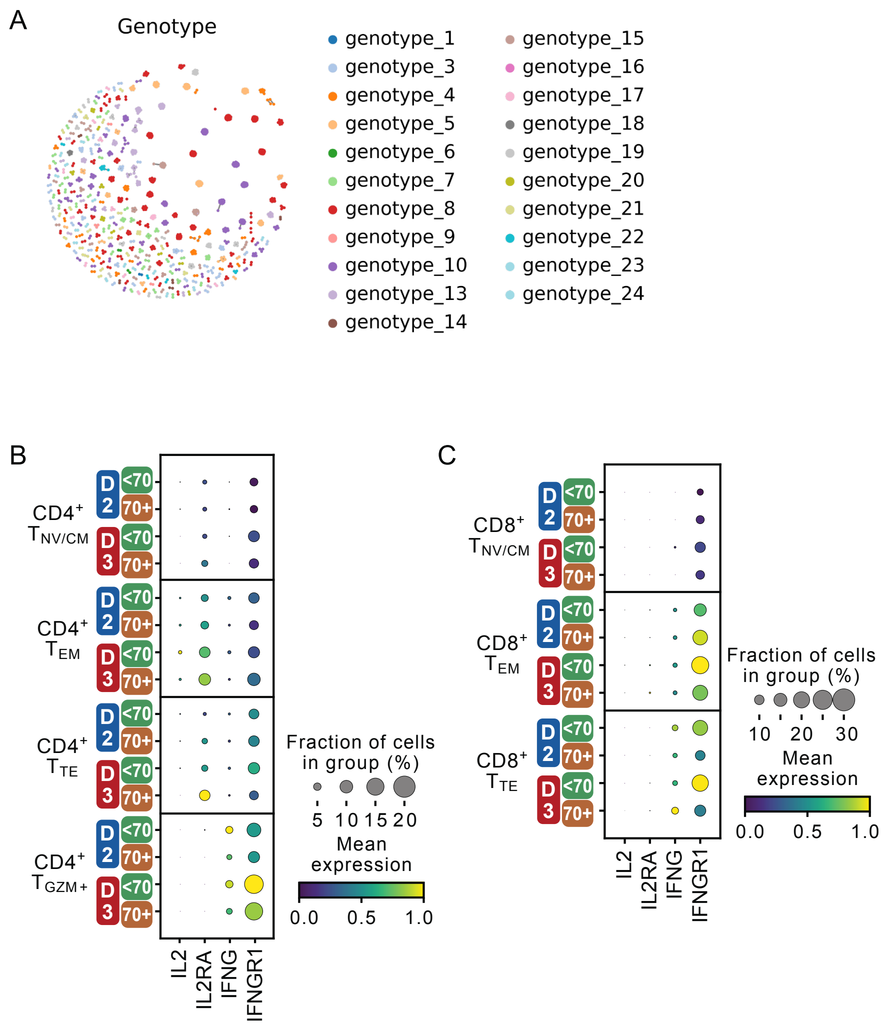

#### Supplementry figure 9

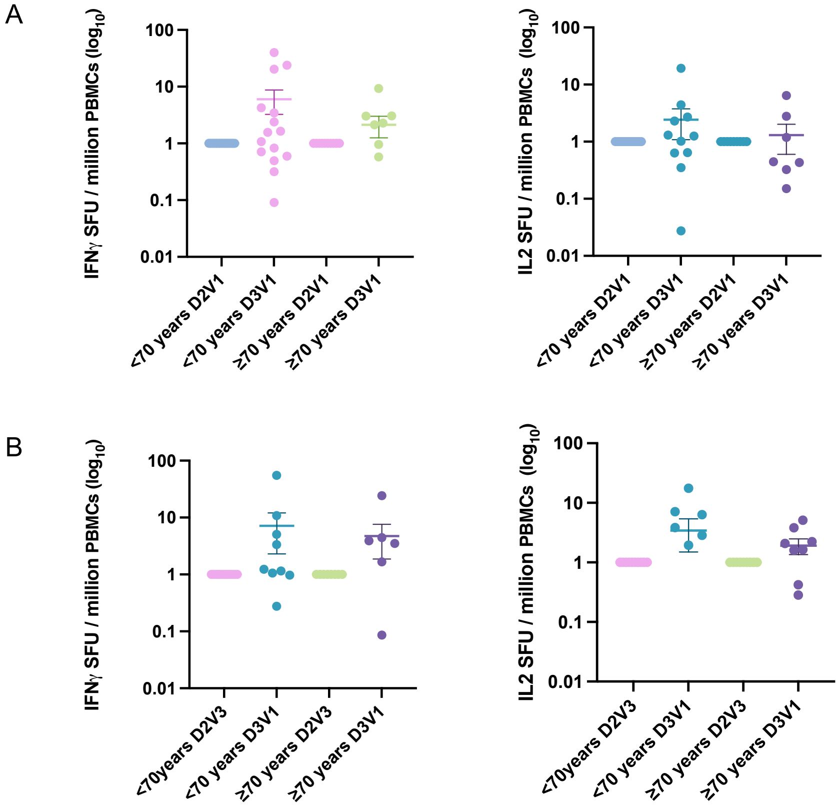

### Supplementary figure 10

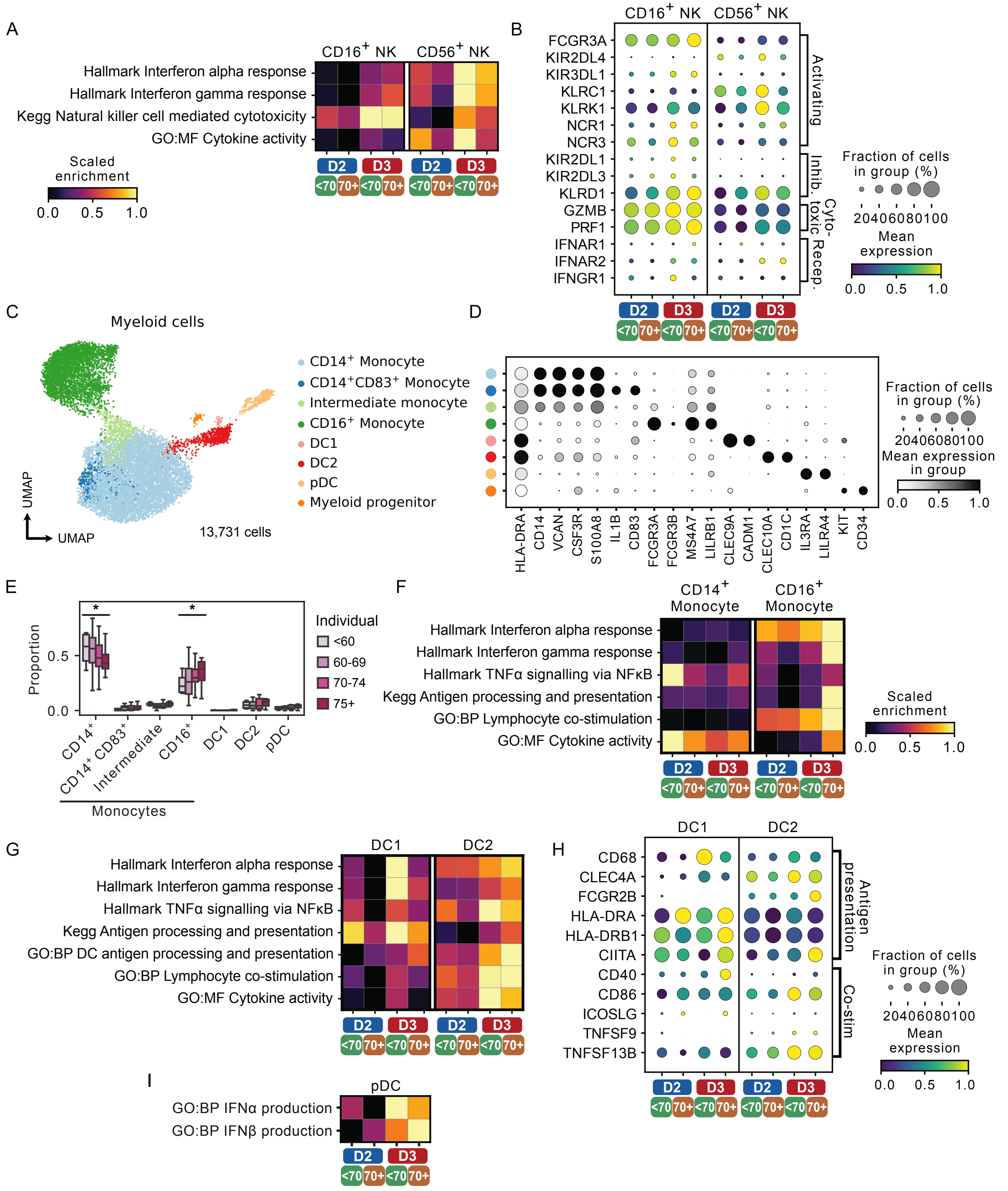
